# Dynamics of gut microbiota and short-chain fatty acids during a cycling Grand Tour are related to exercise performance and modulated by dietary intake

**DOI:** 10.1101/2022.12.03.22282948

**Authors:** Manuel Fernández-Sanjurjo, Javier Fernández, Pablo Martínez-Camblor, Manuel Rodríguez-Alonso, Raquel Ortolano-Ríos, Paola Pinto-Hernández, Juan Castilla-Silgado, Almudena Coto-Vilcapoma, Lorena Ruiz, Claudio J Villar-Granja, Cristina Tomás-Zapico, Abelardo Margolles, Benjamín Fernández-García, Eduardo Iglesias-Gutiérrez, Felipe Lombó

**Author notes:** **Correspondence to:** Benjamín Fernández-García, MD, PhD, Department of Morphology and Cell Biology, School of Medicine and Health Sciences, University of Oviedo, Avenida Julián Clavería, 6. 33006 Oviedo, Asturias, Spain. MFS and JF are joint first authors. EIG and FL are joint senior authors.

## Abstract

**Objective:** The aim of this study was to analyse the dynamics of faecal microbiota composition and short-chain fatty acids (SCFAs) content of professional cyclists over a Grand Tour, and their relationship with performance and dietary intake.

**Design:** 16 professional cyclists competing in La Vuelta 2019 were recruited. Faecal samples were collected at four time points: The day before the first stage (A); After 9 stages (B); After 15 stages (C); On the last stage (D). Faecal microbiota populations and SCFAs content were analysed using 16S rRNA sequencing and gas chromatography, respectively.

**Results:** A principal component analysis (PCA) followed by Generalized Estimating Equation (GEE) models were carried out to explore the dynamics of microbiota and SCFAs, and its relationship with performance. *Bifidobacteriaceae, Coriobacteriaceae, Erysipelotrichaceae*, and *Sutterellaceae* dynamics showed a strong final performance predictive value (R^2^=0.83, ranking and R^2^=0.81, accumulated time). Positive correlations were observed between *Coriobacteriaceae* and acetate and isovalerate and *Bifidobacteriaceae* and isobutyrate. No relationship was observed between SCFAs and performance. The abundance of *Bifidobacteriaceae, Coriobacteriaceae*, and *Erysipelotrichaceae* at the beginning of La Vuelta was directly related to the previous intake of a complex-carbohydrate-rich food, potatoes, while during the competition the abundance of *Bifidobacteriaceae* was negatively affected by the intake of simple carbohydrates from supplements.

**Conclusion:** An ecological perspective represents more realistically the relationship between gut microbiota composition and performance, compared to single-taxon approaches. The composition and periodisation of diet and supplementation during a Grand Tour, particularly carbohydrates, could be designed to modulate gut microbiota composition that allow better performance.

**KEY MESSAGES:** *What is already known on this topic:* Gut microbiota composition and metabolic activity change in response to acute exercise and training and can directly influence physical performance. However, the effect of a cycling stage race, which entails repeated and continuous days of competition, with extreme physiological and metabolic demands, has not been analysed.

*What this study adds:* Our results show that microbiota dynamics during a Grand Tour involve many taxa and allow performance prediction. Those dynamics are related to dietary intake, both prior to and during competition.

*How this study might affect research, practice or policy:* Our results demonstrate that limiting the relationship between gut microbiota composition and sports performance to a single taxon or metabolite is a reductionist approach that does not reflect the complexity of the microbial ecosystem. It may help to optimize the composition and periodisation of diet and supplementation based on gut microbiota modulation to optimize performance.

## INTRODUCTION

Gut microbiota composition influences the risk of highly prevalent pathologies, such as cardiovascular and metabolic diseases or cancer.^1 2^ Regular exercise is a protective factor against these pathologies, although the exact mechanisms through which this effect takes place are diverse and yet not well defined.^3^ In this context, interest in the modulatory role of exercise on gut microbiota has emerged in recent years.^4 5^ Regular exercise has been described as a factor modifying both the diversity and the relative abundance of certain bacterial phyla or families, although the few studies that have been carried out on this show important methodological differences. The main divergence between studies lie in the species used (human, rat, mouse); the type of sample in which the microbiota is analysed (faeces and/or caecal content); the presence or absence of dietary control; the frequency, duration, and intensity of exercise interventions (acute exercise or training);^6-8^ and exercise type (endurance, resistance, concurrent training, etc.).^9^

Some authors have described that gut microbiota composition and metabolic activity directly influence physical performance.^10-12^ This puts the focus on the use of sports supplements that could modulate the composition and/or the metabolic activity of gut microbiota in order to improve performance.^13^ Although it has been described that gut microbiota composition can be modified with the use of different supplements (probiotics, prebiotics, proteins, antioxidants, branched-chain amino acids, caffeine, etc.),^14^ the lack of solid information on how exercise modifies the composition of the gut microbiota, especially in high-level athletes during competition, makes this supplementation imprecise and potentially ineffective.

To our knowledge, the effect of a cycling stage race on the gut microbiota composition and metabolic activity of professional cyclists has not been analysed. Grand Tours involve repeated and continuous days of competition, which entail extreme physiological and metabolic demands and responses.^15 16^ Therefore, studying the composition of the intestinal microbiota and the products of its metabolic activity in this context can help both to understand its potential modulatory role in the response to exercise and to optimize and personalize the use of supplements.

Thus, the aim of this study was to analyse the faecal microbiota composition and short-chain fatty acids (SCFAs) content of professional cyclists over the three weeks of a Grand Tour, La Vuelta 2019, and its relationship with performance indicators, dietary intake, and supplement use.

## SUBJECTS AND METHODS

### Subjects

16 professional cyclists from two cycling teams among those competing in La Vuelta 2019 were contacted to participate in this study. Both teams were ranked in the top 10 of the 2019 UCI World Ranking, and both finished in the top 5 in the team classification at La Vuelta 2019. All the cyclists accepted to participate and signed an informed consent form. At the time of recruitment, and at least in the previous month, none of them was using any antibiotic or other pharmacological treatment that could interfere with the intestinal microbiota and that would have constituted a cause for exclusion. During the study, one of the participants suffered a fall that forced his withdrawal from competition. Thus, the total number of subjects who completed the study was 15 cyclists.

Both the study design and the informed consent were reviewed and authorized by the corresponding Ethics Committee (Ref.: 238/19).

### Anthropometric measurements

Body mass and height were measured using a medical scale with a measuring rod (Seca, model 704s; precision: 0.1 kg for weight and 0.1 cm for height; Hamburg, Germany), at the beginning and the end of the competition.

### Collection of faecal samples

Faecal samples were collected at four time points: On the morning of the day before the first stage of La Vuelta 2019 (A); On the morning of the first rest day (after 9 stages) (B); On the morning of the second rest day (after 15 stages) (C); and on the morning of the last day of the race (after 20 stages) (D).

Sampling was performed according to established protocols to avoid cross-contamination using Fecal Nucleic Acid Collection and Preservation Tubes (Norgen Biotek Corp., Thorold, Canada) to preserve the nucleic acids in perfect condition for subsequent analysis.

### Analysis of gut microbiota populations using 16S rRNA sequencing

From 200 mg of faecal samples, genomic DNA (gDNA) was extracted using the E.Z.N.A.® DNA extraction kit (Omega Bio-Tek Ref. D4015-02, GA, USA), obtaining 200 µl of gDNA. The gDNA samples were quantified using the BioPhotometer® (Eppendorf, Hamburg, Germany) and their concentrations diluted and standardized to 6 ng/µl.

These samples were used for PCR amplification using the Ion 16™ Metagenomic kit (Thermo Fischer Scientific, MA, USA). The PCR amplification products were used to create a genomic library using the Ion Plus Fragment Library kit AB library Builder™ System (Cat. No.4477597, Thermo Fischer Scientific, MA, USA). Finally, the Ion Xpress™ Barcode adapters 1-96 kit (Cat. No. 4474517, Thermo Fischer Scientific, MA, USA) were added to each sample for subsequent sequencing.

Sequencing was performed using the ION PGM™ Hi-Q™ Sequencing kit (Cat. No. A25592, Thermo Fischer Scientific, MA, USA) on the ION PGM™ system. The chip used was the 318™ v2 (Cat. No. 4,484,355, Thermo Fischer Scientific, MA, USA). The total number of reads was always greater than 110,000 per sample.

The consensus table for each sequencing run was downloaded from ION Reporter 5.6 software. This table includes the percentages for each taxonomic level and was used to compare frequencies between individuals. Taxonomic adscription down to the species level was performed using QIIME 2 (v.2017.6.0). The reference libraries used were the Curated MicroSEQ™ 16S Reference Library v2013.1 and Greengenes v13.5. Those phyla, families, or species that did not reach 0.1% of relative abundance at any of the sampling points were eliminated for further analysis. From the sequencing information generated, alpha diversity indices, such as Shannon diversity index and Simpson richness and evenness index^17^, were calculated using the QIIME 2 platform. All raw metagenomics data have been deposited at NCBI SRA database (Accession number: <u>PRJNA645285</u>).

### Short chain fatty acids analysis

The SCFAs acetate, propionate, isobutyrate, butyrate, isovalerate, and valerate were determined in faeces through gas chromatography (GC), using a system composed of a 6890NGC injection module (Agilent Technologies Inc., Palo Alto, USA) connected to a flame injection detector (FID) and a mass spectrometry (MS) 5973N detector (Agilent), as previously described. Briefly, cell free-supernatants (100□μl) from faecal homogenates, were mixed with 450□μl methanol, 50□μl internal standard solution (2-ethylbutyric 1.05□mg/ml), and 50□μl 20% v/v formic acid. Following centrifugation, supernatants of this mixture were used for SCFAs quantification by GC as previously described.^18^

### Performance, fatigue perception and recovery

Three objective parameters were measured as indicators of performance: power output, expressed as the average power-to-weight ratio per stage (W/kg), and position in the overall individual ranking and accumulated time at each sampling point, obtained from the data of the official competition record (https://www.lavuelta.es/es/clasificaciones). On the other hand, two subjective parameters were measured: the Rating of Perceived Exertion (RPE) at the end of La Vuelta, as an indicator of fatigue, and the Total Quality Recovery (TQR) scale, both at the beginning and at the end of the competition, as an indicator of recovery. For the analysis of the RPE, the Borg scale 6-20 ^19^ was used, being 6 “very very light” and 20 “very very hard”. Perception of recovery was evaluated using a 6-20 scale,^20^ being 6 “very, very poor recovery” and 20 “very, very good recovery”.

### Dietary assessment and recording of probiotic supplement use

Food intake and supplement use recording before and during La Vuelta was only provided by the medical staff of one of the teams (n=8).

A retrospective analysis of dietary intake during the month preceding the start of La Vuelta was carried out using a validated food frequency questionnaire,^21^ which was modified based on a quantitative study on food consumption of elite cyclists during training and competition^22^ and including specific sports foods.^23^ The food items added to the original questionnaire were: wholegrain cereals and derivatives (bread, pasta, rice, breakfast cereals, cookies, etc.), olive oil, other vegetable oils (sunflower, corn, canola, etc.), butter, margarine, and fermented dairy products other than yogurt (kefir, koumiss, etc.). Moreover, items corresponding to milk and dairy products were itemized as: whole, semi-skimmed, and skimmed milk, and full-fat and low-fat yogurt and fermented dairy products. Regarding sports foods, the use of carbohydrate (CHO) drinks, CHO/protein bars, and CHO/caffeinated gels was also included.

The same questionnaire was used to assess food intake during competition. The cyclists were asked to record their intake two days before B sampling point. In agreement with the teams’ medical staff, the cyclists were asked to repeat the same dietary pattern two days before C and D sampling points to minimize the influence of this variable. The use and composition of probiotic supplements throughout the competition was also recorded from each cyclist by the medical staff, as well as any antibiotic treatment.

### Statistical Analysis

Pearson’s correlation coefficients were used for studying the correlation between the variables. Comparison between groups were based on one-way ANOVA or Kruskal-Wallis, and repeated measured ANOVA or Friedman test as appropriate. A Principal Component Analysis (PCA) was performed at point A for both microbiota composition at family level (lowest taxonomic category for which the sequencing technique used allows the identification of 100% of the taxa) and SCFA. PCA had two-fold objective i) a dimensionality reduction, ii) identify latent components of the gut microbiota and SCFAs. Using varimax rotation, we extract components explaining, at least, the 50% of the total variance. The model was applied at the other sampling points (B-D). Finally, in order to consider the effects of repeated measures on the same subjects, Generalized Estimating Equation (GEE) models with unstructured variance-covariance matrix structure were used for modelling the relationship between the performance indicators and the microbiota and SCFA components. Standard linear regression models were used for developing prediction models at the end of the race. A leave-one-out cross-validation procedure was used for reducing the overfitting in its accuracy evaluation. All reported p-values were two-sided and those below 0.05 were considered statistically significant. R.4.01 (www.r-project.org) was used for statistical analysis.

## RESULTS

### Gut microbiota and SCFAs composition varies throughout La Vuelta

We studied 15 professional cyclists at four different time-points during La Vuelta 2019, which characteristics are shown in Table 1. Considering all sampling points, faecal 16S metagenomic dataset yielded a total of 8 phyla, 42 families, 45 genera, and 75 species (Supplementary Tables S1, phyla; S2, families; and S3, genera and species). The sequencing technique used allowed the identification of 100% of the taxa from phylum to family levels. Shannon and Simpson indexes were calculated at family level, and no significant differences were observed between sampling points (A-D), being 3.3±0.4, 3.1±0.4, 3.1±0.4, and 3.2±0.3 (p=0.405) and 0.81±0.06, 0.81±0.06, 0.79±0.06, and 0.79±0.05 (p=0.433), respectively (Supplementary Figure S1).

**Table 1.**
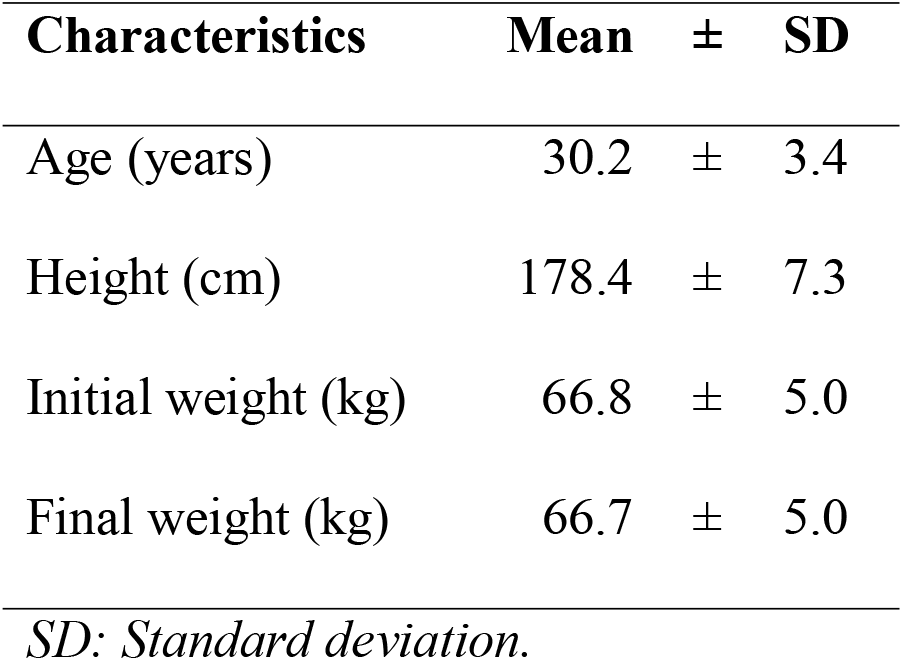
Physical characteristics of the subjects (n=15)

Were observed changes in the abundance of certain phyla and families between sampling points. Thus, the relative abundance of phylum Actinobacteria increased significantly (p=0.000331) from A (0.6±1.6%) to B (3.0±3.4%) (Figure 1). As the scenario was more complex at the family level, we decided to perform a PCA in order to understand gut microbiota dynamics throughout La Vuelta. This PCA was defined at point A and then applied at the other sampling points (B-D). Three principal components (PC1, PC2, and PC3) explained 50% of the variance in family abundance at A (Figure 2A). The dynamics of this model is shown in Figure 2B. The main families included in these PCs (absolute value of the load larger than 0.5) and their specific response are shown on Figure 2C for PC1, 2D and 2E for PC2, and 2F for PC3.

**Figure 1.**
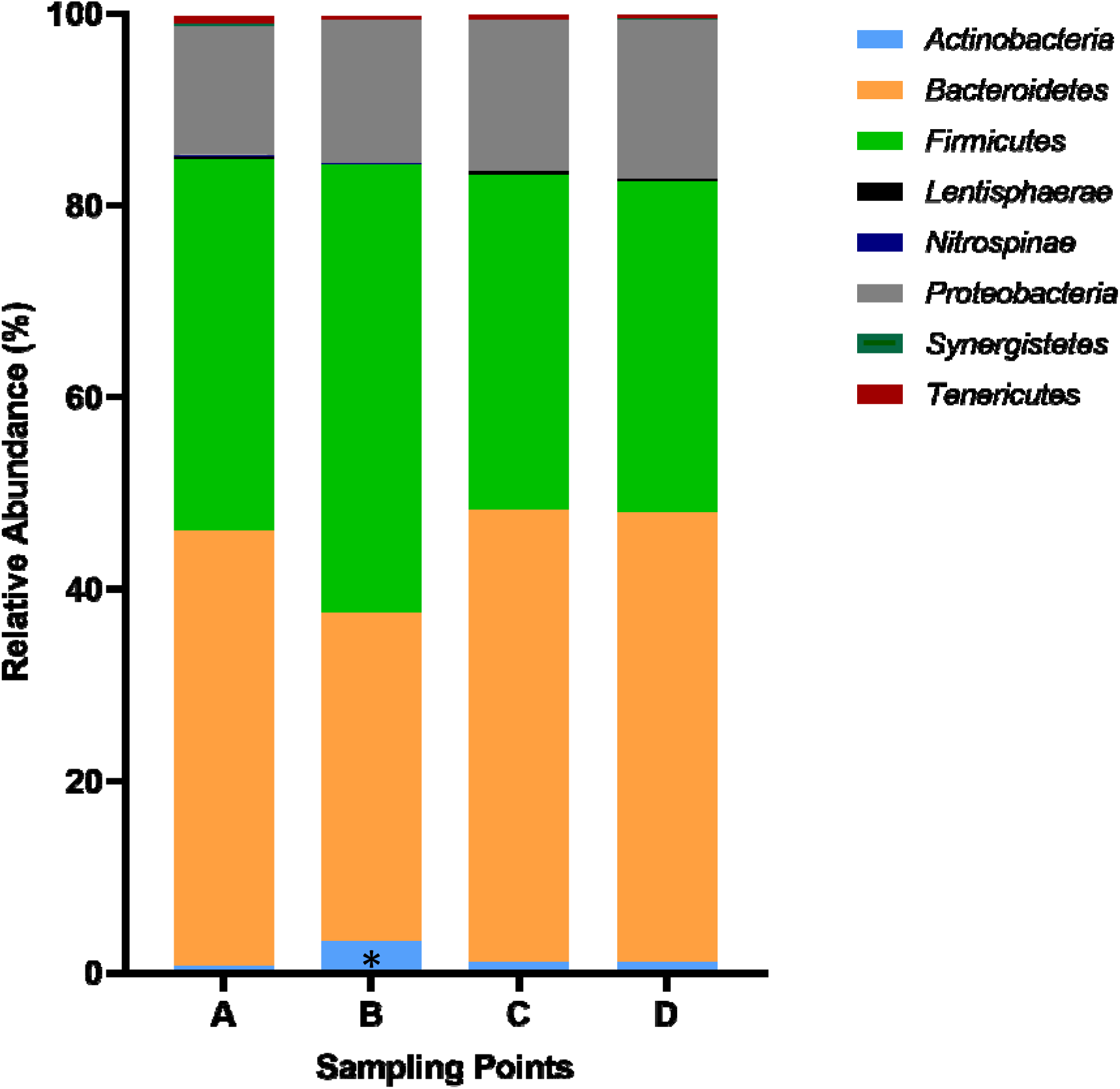
Relative abundance of phyla during La Vuelta. A: Sampling point immediately before the first stage; B: Sampling point after 9 stages; C: Sampling point after 16 stages; D: Sampling point after 20 stages. *: Statistically significant difference between *Actinobacteria* A vs. B (p=0.000331).

**Figure 2.**
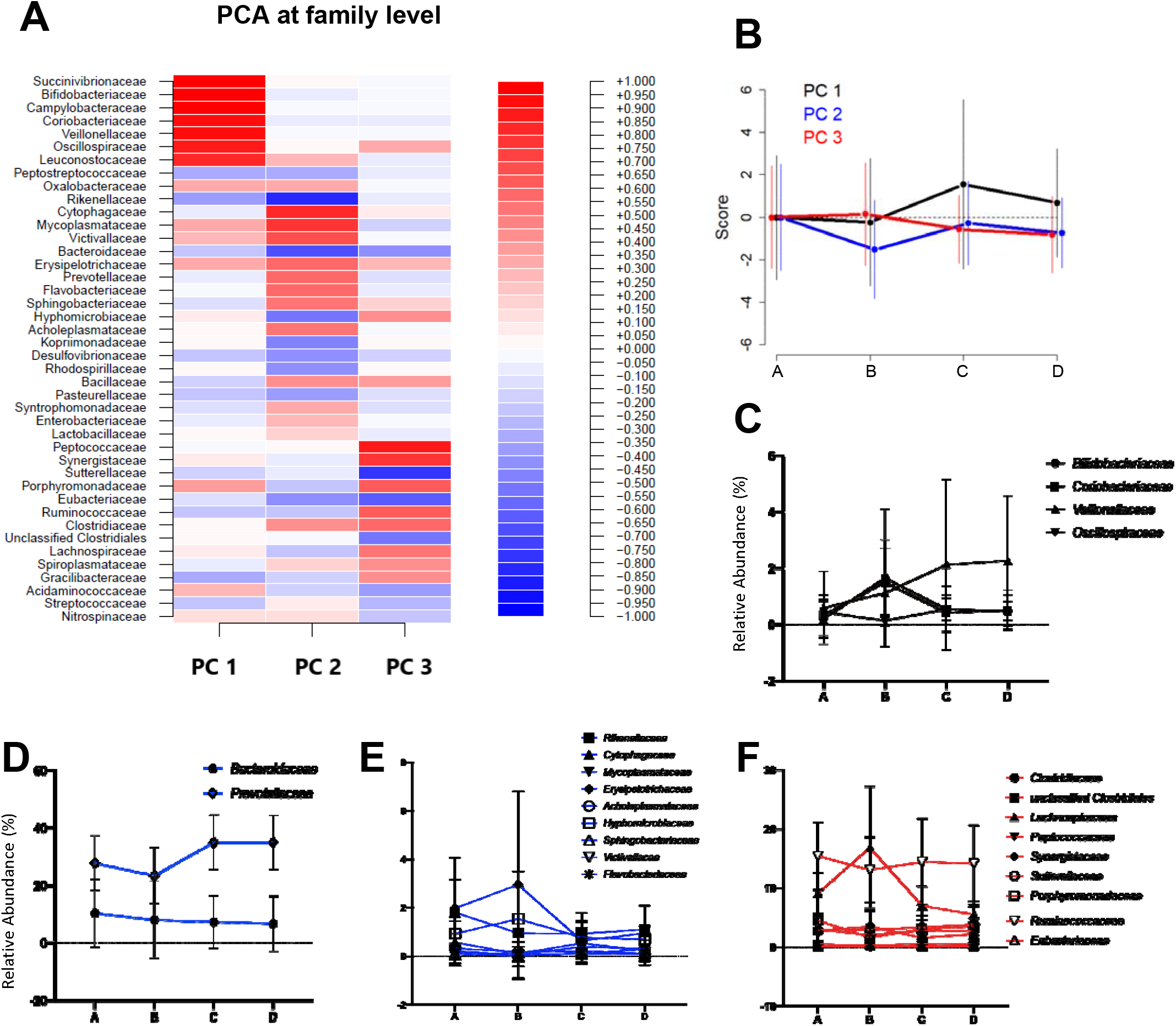
Microbiota dynamics during La Vuelta. **A**. Heatmap of the principal component analysis (PCA) at family level, including the coefficients for each family in the different PCs identified. **B**. Dynamics of the PCs in the different sampling points, A-D. It shows the mean and standard deviation of the score obtained by applying the model at each sampling point. **C**. Relative abundance of the families with coefficients <-0.5 or >0.5 in the PC1 at the different sampling points, A-D. **D**. Relative abundance at the different sampling points, A-D, of the families with coefficients <-0.5 or >0.5 in the PC2 and relative abundances >5%. **E**. Relative abundance at the different sampling points, A-D, of the families with coefficients <-0.5 or >0.5 in the PC2 and relative abundances <5%. **F**. Relative abundance of the families with coefficients <-0.5 or >0.5 in the PC3 at the different sampling points, A-D. A: sampling point immediately before the first stage; B: sampling point after 9 stages; C: sampling point after 16 stages; D: sampling point after 20 stages.

A PCA was also carried out following the same approach to analyse the dynamics of faecal SCFAs during La Vuelta (Figure 3A and 3B). One principal component (PC1) explained the 70% of the variance at A, with all SCFAs participating with a positive score. The specific response of each SCFA is shown on Figure 3C.

**Figure 3.**
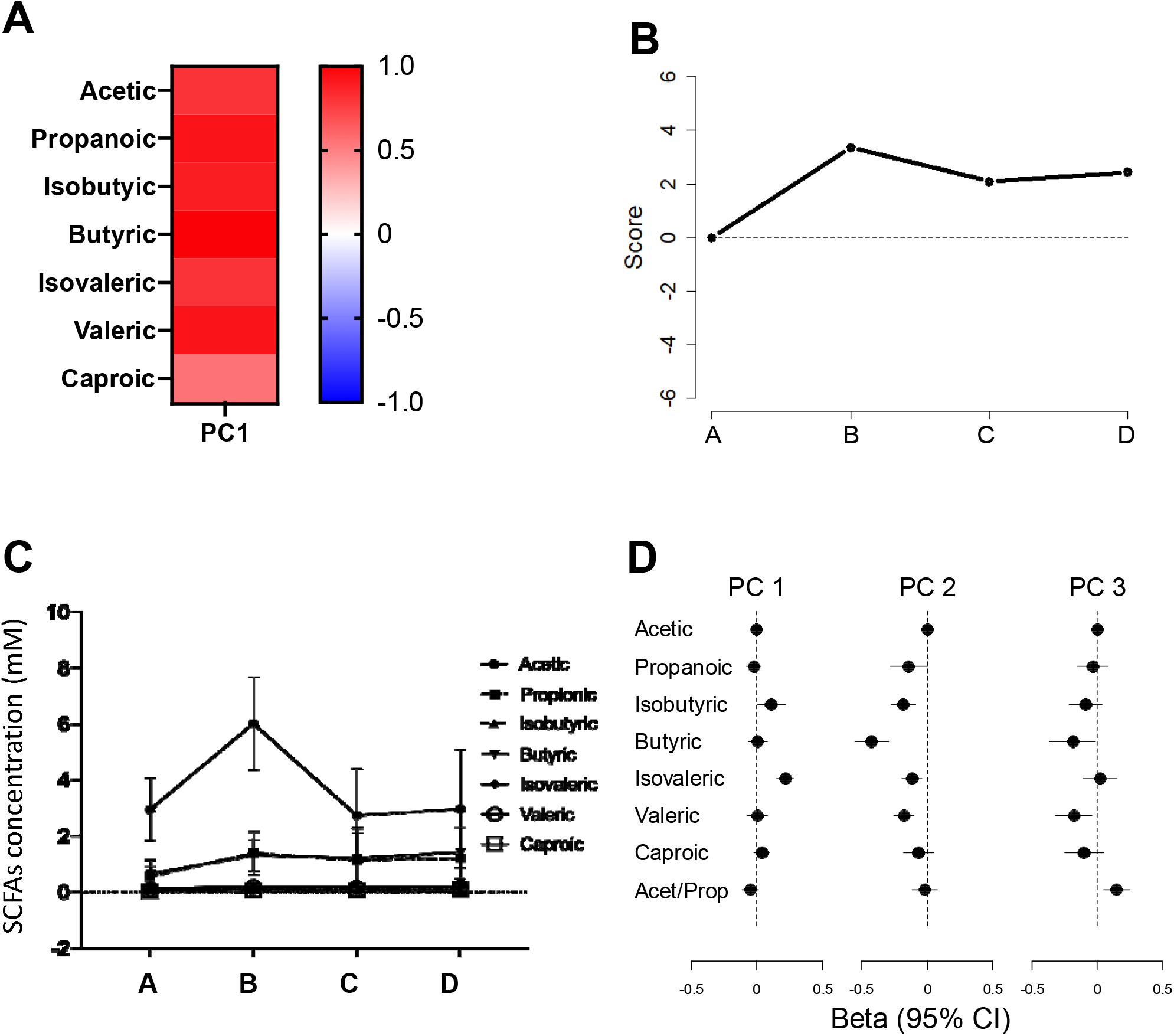
Short-chain fatty acids (SCFA) dynamics during La Vuelta. **A**: Heatmap of the principal component analysis (PCA), including the coefficients for each SCFA in the PC identified. **B**. Dynamics of the PC in the different sampling points, A-D. It shows the mean and standard deviation of the score obtained by applying the model at each sampling point. **C**. Relative abundance of the SCFAs at the different sampling points, A-D. **D**. Forest plot showing the global relationship between the principal components identified for microbiota and SCFAs. The dots represent the estimated effect (beta) and the lines the 95% confidence interval. A: Sampling point immediately before the first stage; B: Sampling point after 9 stages; C: Sampling point after 16 stages; D: Sampling point after 20 stages.

To explore the relationship between faecal microbiota composition and SCFAs concentration, GEE models were used. A positive relationship was observed between microbiota PC1 and isovaleric (p<0.0001) and isobutyric (p=0.043) acids. Besides, microbiota PC2 and PC3 showed a negative relationship with SCFAs. Specifically, PC2 with propionic acid (p=0.048), isobutyric (p<0.0001), butyric (p<0.0001), isovaleric (p=0.002), and valeric (p<0.0001) acids; and PC3 with butyric (p=0.034) and valeric (p=0.011) acids. A forest plot with these relationships is shown in Figure 3D. Considering that an increase in SCFA faecal content was observed at sampling point B (Figure 3B), a correlation analysis was performed at this sampling point between specific SCFAs concentration, and the abundance of those families related to performance according to GEE models, as well as some of its genera and species. Positive correlations were observed at this sampling point for *Coriobacteriaceae* and its species *Collinsella aerofaciens* with acetic (Pearson correlation coefficient of 0.530, 95% CI: 0.024-0.820, p=0.042; and 0.698, 95% CI: 0.290-0.892, p=0.004) and isovaleric acids (Pearson correlation coefficient of 0.664, 95% CI: 0.230-0.878, p=0.007; and 0.549, 95% CI: 0.052-0.829, p=0.034) and for *Bifidobacteriaceae* and its genus *Bifidobacterium* with isobutyric acid (Pearson correlation coefficient of 0.682, 95% CI: 0.262-0.885, p=0.005; and 0.682, 95% CI: 0.262-0.885, p=0.005) (Supplementary Figure S2). No correlation was observed between *Sutterellaceae* abundance and SCFA concentration at this sampling point.

Interestingly, sampling point B was also relevant for performance. A strong significant positive correlation was observed between the classification at points B and D (Pearson correlation coefficient of 0.951, 95% CI: 0.855-0.984; p<0.0001) (Figure 4A). Furthermore, we observed that those cyclists performing a higher average power-to-weight ratio per stage from A to B obtained better results in terms of better final ranking (Pearson correlation coefficient of -0.781, 95% CI: (−0.927)-(−0.427); p=0.000981) (Figure 4B) and lower accumulated time at the end of La Vuelta (Pearson correlation coefficient of -0.818, 95% CI: (−0.941)-(−0.508); p=0.000347) (Figure 4C). This correlation was not observed from B to C or from C to D, although we found it between the average power-to-weight ratio per stage throughout La Vuelta and both final ranking (Pearson correlation coefficient of -0.627, 95% CI: (−0.875)-(−0.116); p=0.022) (Figure 4D) and final accumulated time (Pearson correlation coefficient of -0.642, 95% CI: (−0.881)-(−0.141); p=0.018) (Figure 4E).

**Figure 4.**
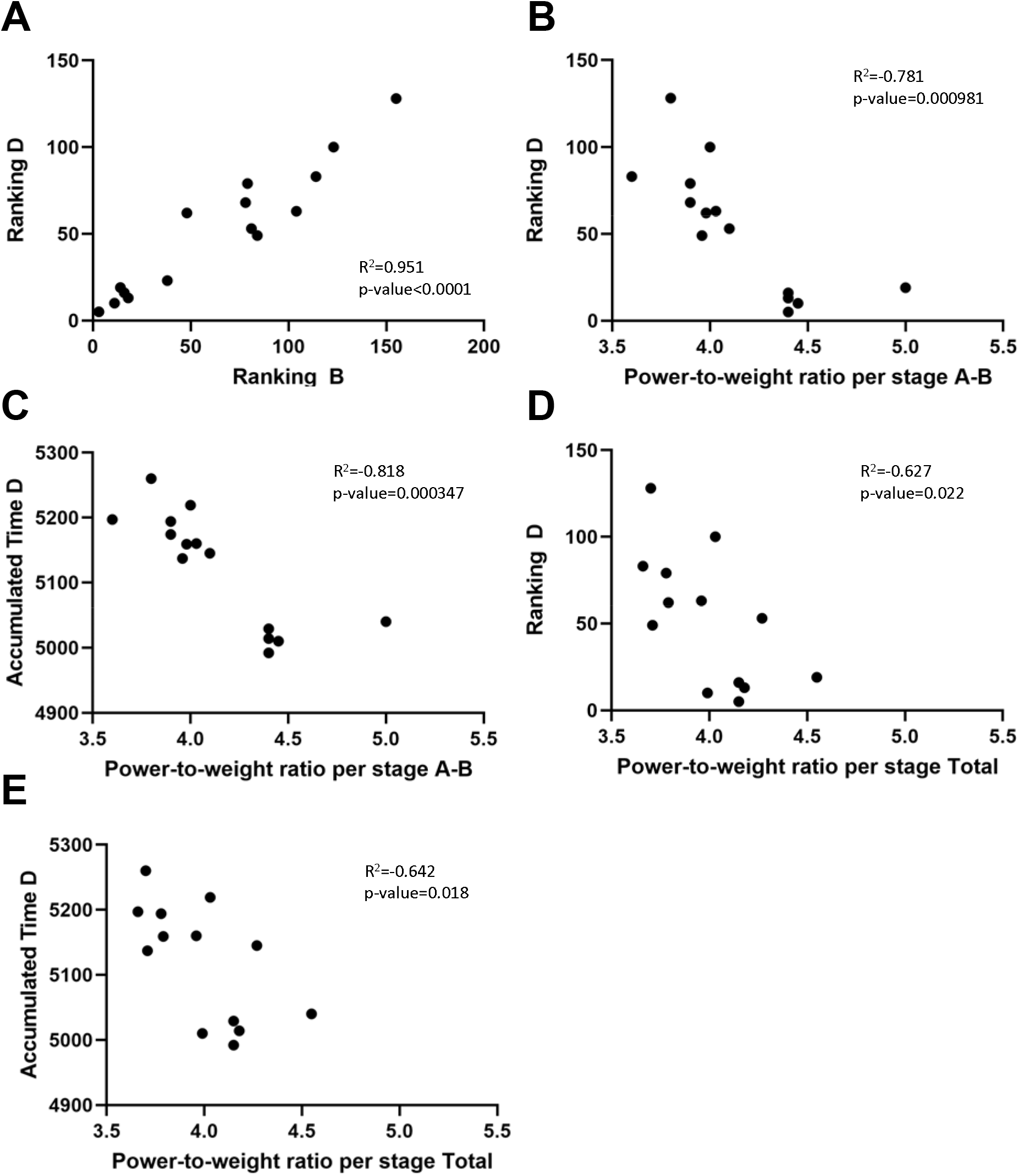
Performance parameters during La Vuelta. **A**: Correlation analysis between ranking at points B and D. **B**. Correlation analysis between final ranking (timepoint D) and power-to-weight ratio per stage between timepoints A and B. **C**. Correlation analysis between final accumulated time and power-to-weight ratio per stage between timepoints A and B. **D**. Correlation analysis between final ranking (timepoint D) and power-to-weight ratio per stage between timepoints A and D. **C**. Correlation analysis between Final accumulated time and power-to-weight ratio per stage between timepoints A and D. A: Sampling point immediately before the first stage; B: Sampling point after 9 stages; C: Sampling point after 16 stages; D: Sampling point after 20 stages.

### Gut microbiota dynamics during La Vuelta predicts performance

In addition to analysing changes in gut microbiota composition and SCFAs concentration throughout La Vuelta, we recorded performance, expressed as the position in the overall individual ranking and as accumulated time (Table 2). Based on a linear regression model, we explored the relationship between gut microbiota PCA models and accumulated time and ranking at the end of La Vuelta (Figure 5). Using PC1, PC2 and PC3 together, a strong performance predictive value was observed, both for accumulated time (Pearson correlation coefficient of 0.83, p=0.0004348, Figure 5A) and for ranking (Pearson correlation coefficient of 0.81, p=0.0006532, Figure 5B). Focusing specifically on each PC, we observed that those cyclists with high levels of PC2 at A, high levels of PC1 and PC2 at B, and low levels of PC2 and high levels of PC3 at C performed better, obtaining a higher final ranking, i.e., less accumulated time. Deepening this analysis, the specific representative families within each PC follow these trends are *Bifidobacteriaceae* and *Coriobacteriaceae* for PC1, *Erysipelotrichaceae* for PC2, and *Sutterellaceae* for PC3 (Figures 2C, E and F). Interestingly, the models provided no relationship between SCFAs and performance. Furthermore, no relationship was observed between the dynamics of microbiota and SCFA with other performance, recovery, and fatigue parameters, such as the average power-to-weight ratio per stage, TQR or RPE.

**Table 2.** Fatigue and recovery perception and performance parameters at different sampling points throughout competition (n=15)

| Parameter | Mean | ± | SD |
| --- | --- | --- | --- |
| TQR, A | 17.7 | ± | 1.7 |
| TQR, D | 12.7 | ± | 7.3 * |
| RPE, D | 16.3 | ± | 1.5 |
| Classification position, B | 67 | ± | 47 † |
| Classification position, C | 56 | ± | 40 |
| Classification position, D | 51 | ± | 37 |
| Accumulated time (min), B | 2172.2 | ± | 38.2 |
| Accumulated time (min), C | 3829.3 | ± | 63.7 |
| Accumulated time (min), D | 5119.5 | ± | 86.7 |
| Average power-to-weight ratio per stage ($W \cdot kg^{-1}$), A-B | 4.2 | ± | 0.4 ‡ |
| Average power-to-weight ratio per stage ($W \cdot kg^{-1}$), B-C | 3.9 | ± | 0.3 |
| Average power-to-weight ratio per stage ($W \cdot kg^{-1}$), C-D | 3.8 | ± | 0.4 |
| Average power-to-weight ratio per stage ($W \cdot kg^{-1}$), Total | 4.0 | ± | 0.3 |

*SD: Standard deviation. A: Sampling point before the first stage; B: Sampling point after 9 stages; C: Sampling point after 16 stages; D: Sampling point after 20 stages. \*: Statistically significant difference between TQR A vs. D ( $p=0.000744$ ); †: Statistically significant difference between Classification position B vs. D ( $p=0.017$ ); ‡: Statistically significant difference between Average power-to-weight ratio per stage A-B vs. B-C ( $p=0.036$ ) and C-D ( $p=0.006$ ).*

**Figure 5.**
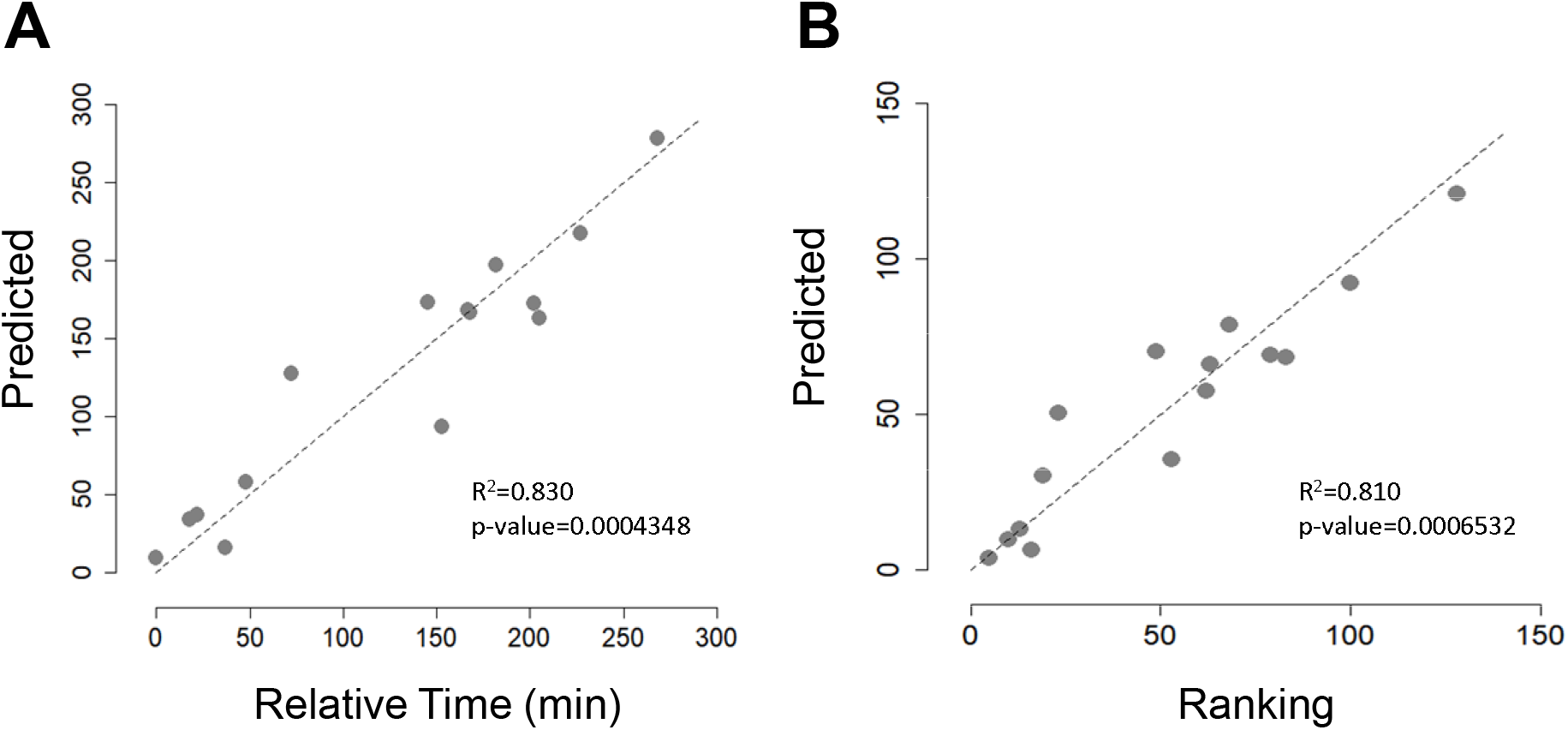
Performance prediction based on microbiota components. **A**: Correlation analysis between predicted final accumulated time based on GEE models analysis and relative time, measured as the difference in the final accumulated time of each cyclist with respect to that of the best classified of those included in the study. **B**: Correlation analysis between Predicted final ranking based on GEE models analysis and final ranking.

### Dietary intake modifies microbiota composition to modulate performance

Considering that both microbiota composition and physical performance are modulated by diet, food intake and sport supplement use, including probiotics, in the month prior to La Vuelta and during the competition was recorded by means of a food frequency questionnaire. The results are shown in Table 3. A significant increase was observed during La Vuelta in the frequency of consumption of food groups rich in refined CHO, such as pasta (Percent variation=350±382%), rice (177±111%), bread (114±210%), or soft drinks (136±95%), although the intake of these food groups was already high previously. Similarly, the consumption of high-CHO sport supplements increased significantly during competition (CHO drinks: 442±677%; Gels: 533±47%; Energy bars: 288±214%; Sport snacks: 167±145%). The main probiotic food consumed was yoghourt, which includes members of the genus *Lactobacillus* and the species *Streptococcus thermophilus*. The athletes studied did not consume probiotic supplements in the weeks prior to La Vuelta, although all used these supplements daily during the competition. The commercial products used included combinations of the species *Bifidobacterium lactis, Bifodobacterium bifidum, Lactobacillus paracasei, and Lactobacillus acidophilus*.

**Table 3.** Frequency of consumption (portions per week) of foods and sport supplements before and during La Vuelta 2019 (n=8)

| **Food item** | **Before competition**<br>**(portions per week)** | **During competition**<br>**(portions per week)** | **p-value** |
| --- | --- | --- | --- |
| *Supplements* |  |  |  |
| Carbohydrate drinks | 3.2 ± 3.0 | 18.5 ± 14.5 | **0.012** |
| Gels | 0.8 ± 1.5 | 15.9 ± 6.3 | **0.011** |
| Energy bars | 4.4 ± 4.1 | 18.1 ± 8.5 | **0.018** |
| Sport snacks | 12.8 ± 11.1 | 33.1 ± 12.6 | **0.028** |
| Protein bars | 1.1 ± 1.6 | 1.9 ± 2.8 | 0.285 |
| *Foods* |  |  |  |
| Breakfast cereals | 4.6 ± 7.2 | 10.0 ± 11.5 | **0.027** |
| Wholegrain breakfast cereals | 3.1 ± 3.4 | 4.4 ± 5.2 | 0.109 |
| Bread | 11.1 ± 9.1 | 18.4 ± 10.9 | **0.046** |
| Wholegrain bread | 2.0 ± 3.2 | 5.2 ± 6.2 | 0.102 |
| Rice | 3.9 ± 3.4 | 9.0 ± 4.5 | **0.011** |
| Brown rice | 0.4 ± 1.1 | 0.0 ± 0.0 | 0.317 |
| Pasta | 2.5 ± 1.8 | 7.6 ± 2.7 | **0.012** |
| Wholegrain pasta | 0.2 ± 0.7 | 0.0 ± 0.0 | 0.317 |
| Biscuits | 2.5 ± 4.2 | 8.1 ± 6.2 | **0.016** |
| Pastry products | 3.4 ± 7.3 | 4.9 ± 7.4 | 0.109 |
| Chocolate | 3.6 ± 6.9 | 4.8 ± 12.3 | 1.000 |
| Fruits | 12.5 ± 11.0 | 11.4 ± 7.5 | 0.500 |
| Fresh fruit juices | 1.1 ± 2.5 | 2.5 ± 2.9 | 0.141 |
| Commercial fruit juices | 0.4 ± 1.1 | 0.0 ± 0.0 | 0.317 |
| Leafy vegetables | 8.0 ± 4.7 | 9.1 ± 4.9 | 0.396 |
| Other vegetables | 8.8 ± 3.6 | 9.4 ± 4.2 | 0.705 |
| Potatoes | 5.9 ± 6.9 | 5.8 ± 5.3 | 0.344 |
| Legumes | 3.9 ± 4.8 | 5.0 ± 6.6 | 0.343 |
| Nuts | 2.1 ± 2.4 | 3.0 ± 3.3 | 0.414 |
| Whole milk | 3.4 ± 3.2 | 3.6 ± 3.2 | 0.414 |
| Semi-skimmed milk | 1.8 ± 3.2 | 2.6 ± 5.2 | 0.317 |
| Skimmed milk | 0.9 ± 2.5 | 0.9 ± 2.5 | 1.000 |
| Yoghurt | 13.0 ± 13.1 | 10.8 ± 5.9 | 0.865 |
| Other fermented milks | 0.9 ± 2.5 | 0.8 ± 2.1 | 0.317 |
| Fresh cheese | 3.2 ± 3.3 | 5.1 ± 4.7 | 0.102 |
| Mature cheese | 1.1 ± 2.5 | 2.0 ± 4.9 | 0.317 |
| Dairy desserts | 0.5 ± 0.9 | 1.8 ± 2.8 | 0.180 |
| Ice cream | 1.0 ± 2.4 | 0.8 ± 1.2 | 0.655 |
| Lean meat | 4.8 ± 1.7 | 5.1 ± 1.6 | 0.180 |
| Red meat | 2.4 ± 2.3 | 2.8 ± 2.4 | 0.102 |
| Meat products | 0.4 ± 0.5 | 0.2 ± 0.5 | 0.317 |
| Lean fish | 2.1 ± 3.1 | 2.6 ± 3.0 | 0.180 |
| Oily fish | 3.4 ± 2.4 | 3.2 ± 2.6 | 1.000 |
| Shellfish | 0.4 ± 0.7 | 0.2 ± 0.7 | 0.317 |
| Eggs | 13.5 ± 8.2 | 13.5 ± 5.5 | 0.892 |
| Olive oil | 9.8 ± 8.2 | 10.9 ± 10.2 | 0.180 |
| Other vegetable oils | 0.0 ± 0.0 | 0.0 ± 0.0 | 1.000 |
| Butter | 7.1 ± 6.1 | 8.1 ± 8.3 | 0.343 |
| Margarine | 0.0 ± 0.0 | 0.0 ± 0.0 | 1.000 |
| Salty snacks | 1.2 ± 2.4 | 0.9 ± 2.5 | 0.180 |
| Precooked food | 0.0 ± 0.0 | 0.0 ± 0.0 | 1.000 |
| Soft drinks | 1.4 ± 2.1 | 5.2 ± 5.2 | **0.043** |
| Diet drinks | 0.9 ± 2.5 | 0.0 ± 0.0 | 0.317 |
| Wine | 1.2 ± 1.6 | 1.6 ± 1.4 | 0.180 |
| Beer | 1.6 ± 3.5 | 0.8 ± 1.5 | 0.180 |
| Distilled alcoholic beverages | 0.0 ± 0.0 | 0.0 ± 0.0 | 1.000 |

*Data are presented as Mean ± Standard deviation. P-values in bold indicate*
*statistically significant differences ( $p < 0.05$ ) between before and during competition.*

The influence of dietary intake during the month preceding the start of La Vuelta in the abundance at sampling point A of those families more closely related to performance was analysed. A significant positive correlation was observed between *Erysipelotrichaceae* relative abundance and potatoes intake (Pearson correlation coefficient of 0.956, 95% CI: 0.771-0.992; p=0.000203) (Figure 6A). According to the performance prediction analysis presented above, high levels of PC2, where *Erysipelotrichaceae* participates, in A are related to a lower accumulated time at the end of La Vuelta, therefore, better performance.

**Figure 6.**
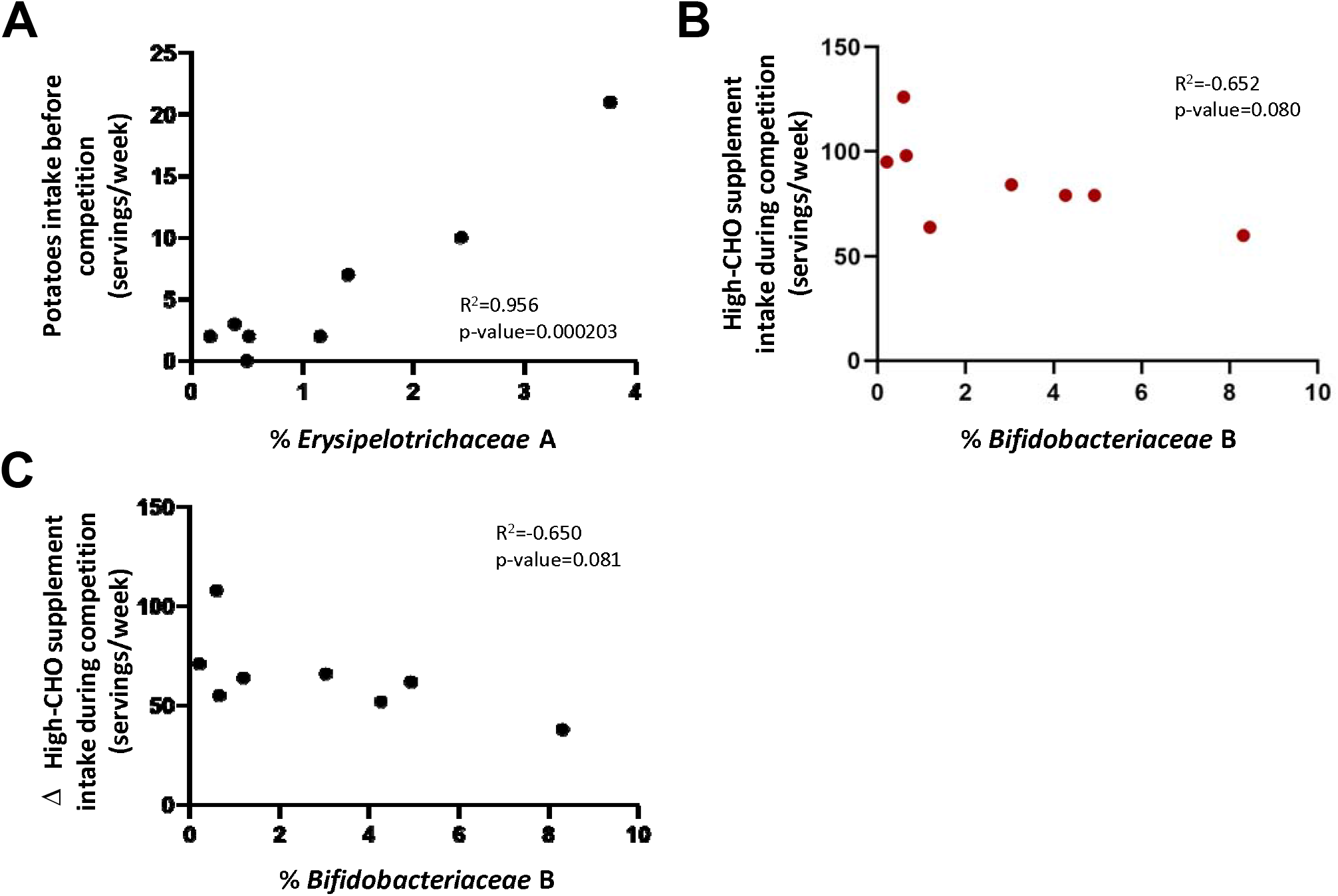
Modulatory effect of diet on microbiota composition before and during La Vuelta. **A**: Correlation between *Erysipelotrichaceae* abundance at timepoint A and potatoes intake before La Vuelta. **B**: Correlation between consumption of supplements high in simple CHO during La Vuelta and *Bifidobacteriaceae* abundance at timepoint B. **C**: Correlation between *Bifidobacteriaceae* abundance at timepoint B and the difference in supplements high in simple CHO consumption during-before La Vuelta.

We also studied this relationship during competition. Those cyclists with the lower frequency of consumption of CHO-rich supplements (CHO drinks, gels, and sport snacks, taken together) during La Vuelta showed the higher relative abundance of the family *Bifidobacteriaceae* at sample point B (Pearson correlation coefficient of -0.652, 95% CI: -0.930-0.097; p=0.080, Figure 6B), where the abundance of this taxon increases considerable regarding A (Figure 2C). Moreover, although the consumption of high-CHO sport supplements increased significantly during competition, as mentioned before, those cyclists with the lower increase in their consumption showed the higher abundance of *Bifidobacteriaceae* family at B (Pearson correlation coefficient of -0.650, 95% CI: -0.929-0.101; p=0.081, Figure 6C). It is worthy to mention that, according to the performance prediction analysis presented above, high levels of PC1, where *Bifidobacteriaceae* participates, in B are related to a lower accumulated time at the end of La Vuelta, therefore, better performance.

The species consumed in both probiotic foods and supplements belonged to the *Bifidobacteriaceae* and *Lactobacillaceae* families. The former being important in PC1 (Figure 2A) and related to performance, as mentioned above. The latter was not relevant in any of the PCs (Figure 2A) and, therefore, did not intervene in the prediction of performance.

## DISCUSSION

This is the first study to analyse changes in faecal microbiota composition and SCFAs concentration during a Grand Tour cycling race. We have also been able to identify those taxa whose timing and magnitude of change during competition show a very close relationship with performance and allow its prediction. Accordingly, this study provides unique information on bacterial dynamics and metabolic activity in response to stress imposed by the accumulation of exercise loads during three weeks of competition and its potential role as performance modulators.

Our results show that the dynamics of the microbiota during La Vuelta involve many taxa, with diverse metabolic activity. Those dynamics result from the demands imposed by exercise and are related to dietary intake, both prior to and during La Vuelta. The mechanisms by which gut microbiota may influence exercise performance have not yet been fully elucidated, although it appears that microbial SCFA production may play a role in muscle energy metabolism, both by being used directly and by affecting the availability of other substrates.^10 24 25^ In fact, well-known families involved in SCFAs production were relevant in the microbiota dynamics during La Vuelta,^26^ such as acetate producers like *Bifidobacteriaceae* and *Coriobacteriaceae*, and diverse propionate and butyrate producers, such as *Oscillospiraceae* and *Veillonellaceae*, from PC1; *Bacteroidaceae, Prevotellaceae* and *Erysipelotrichaceae*, from PC2; and *Lachnospiraceae, Ruminococcaceae, Porphyromonadaceae*, and *Sutterellaceae*, from PC3. Several of these SCFA-producing families allow performance prediction according to the GEE models, such as *Coriobacteriaceae* and *Bifidobacteriaceae*, from PC1, *Erysipelotrichaceae*, from PC2, and *Sutterellaceae* from PC3. These families have previously been described as associated with improved physical performance and exercise-related health status in mouse and human studies.^27 28^ Thus, *Coriobacteriaceae* abundance has been described as a biomarker linking physical exercise with improved health status, probably via diverse catabolic pathways, such as secondary bile acids and aldosterone 18-glucuronide metabolism, involved in sodium homeostasis.^29^ The genus *Bifidobacterium*, main member of the family *Bifidobacteriaceae*, has been associated to improved sport performance and lower plasma levels of parameters associated to fatigue, such as lactate, ammonia, creatine kinase, and inflammatory biomarkers, like pro-inflammatory cytokines, without changes in SCFA production.^30 31^ In addition, *Erysipelotrichaceae* was found to be associated with VO_2_peak, due to its ability to increase butyrate production,^32^ while *Sutterella*, the main genus belonging to the family *Sutterellaceae*, has been positively associated with this parameter through its effect on antioxidant defences^33^. Therefore, despite being well known SCFA producers, the mechanism by which these families are related to performance improvements is not only mediated by their ability to produce SCFAs, but also other metabolites. Interestingly, we have not found a relationship between faecal SCFA content and athletic performance, probably because their faecal content only accounts for about 5% of total SCFA production,^34^ which represents the balance between production, direct uptake by intestinal epithelial cells, and diffusion from the lumen into the portal bloodstream.^35^ Once these colonic SCFAs are absorbed to the portal vein (mainly acetate and propionate) and distributed to the liver and other distant organs, they continue performing important functions. For example, at the muscular level, they contribute to the correct functioning of energy metabolism processes, facilitating the beta-oxidation of fats and the possible recovery of muscle glycogen.^36^ Therefore, we cannot rule out the potential role of SCFAs on performance during La Vuelta, despite the lack of correlation observed with their faecal content. Thus, some members of the *Veillonellaceae* and *Lachnospiraceae* families, well known SCFA producers, have been reported to utilize lactate to synthetize propionate^37^, which highlights their potential value in the context of exercise performance^11 38^. In fact, Scheiman et al.^11^ demonstrated in animal models that serum lactate can reach the intestinal lumen by crossing the intestinal epithelial barrier and that intrarectal infusion of propionate improves performance. In addition, *Lachnospiraceae* is a major producer of butyrate. A positive correlation between butyrate production and VO_2_peak has been described^32^. However, the response dynamics of these families are inconsistent with the performance prediction model obtained in our study. Their role as performance enhancers, if any, does not seem to be related to their abundance, but maybe to their metabolic interaction with other taxa, probably as part of the so-called cross-feeding phenomenon^39^.

Interestingly, although the abundance of *Veillonellaceae* was not related to diet at any sampling point, the relative abundance of *Lachnospiraceae* at the beginning of La Vuelta was directly related to the intake of high-fibre foods (whole-grain foods, vegetables, fruits, potatoes, and legumes, taken together) in the preceding weeks (Pearson correlation coefficient of 0.919, 95% CI: 0.608-0.985; p=0.001, Supplementary Figure S3), as previously described in non-athletic population.^40^ However, this relationship was not observed during competition at any point, even though consumption of fibre-rich foods did not change significantly (46.9±23.6 vs. 53.2±24.3 servings per week; p=0.401). This suggests that, although dietary fibre has a clear modulatory effect on the abundance of *Lachnospiraceae* family, this is less important in the presence of repeated competitive loads, when fibre intake is maintained. It would remain to be elucidated what would happen with respect to the relationship of this family to performance if the intake of fibre-rich foods were modified in a targeted manner at specific times during competition. Similarly, the relative abundance of *Erysipelotrichaceae* at the beginning of La Vuelta was directly related with the intake of complex CHO food sources, although this relationship was not observed during competition. According to our performance prediction model, the higher the relative abundance of this family at A, the better for performance. Therefore, in this case, the modulatory role of the pre-competition diet has an impact on performance. Regarding the effect of food intake during competition on the abundance of other bacterial families related to performance, we have found that *Bifidobacteriaceae* is unable to progress in the gut during competition under high intakes of fast digesting CHO from supplements (glucose, fructose, glucose polymers) or when drastically increasing their consumption habits of these types of supplements. This last idea is related to the concept of “gut training” for athletes proposed by Jeukendrup^41^, according to which the adaptations that allow greater intestinal and metabolic utilization of CHO are greater when a diet high in CHO is regularly followed. Our data show that the dynamics of microbiota composition during competition is sensitive to dietary changes that have an impact on sports performance, suggesting that this may be another element to take into account within this “gut training” paradigm.

These results are relevant since they highlight the potential of diet, particularly simple and complex CHO, to mediate or modify performance through the modulation of gut microbiota composition before and during competition. Future strategies to enhance the performance of high-level athletes could include the selective growth of specific targeted bacterial taxa, using *à la carte* prebiotics, instead of the current ineffective probiotic supplementation, which usually cannot maintain stable populations in the gut ecosystem. In fact, we have been unable to detect any of the strains included in the probiotic supplements or in the probiotic foods used. Therefore, although the cyclists were taking probiotic supplements containing bifidobacteria during competition, the relationship between bifidobacteria abundance and performance appears to be modulated by diet in a way that is apparently more relevant to performance than the use of probiotic supplements.

To conclude, the relationship of gut microbiota with sports performance is complex and not determined by single taxa or single metabolites, not even SCFAs, as suggested by certain authors.^11 42^ An ecological perspective seems to represent a more realistic approach to the relationship between the composition and the metabolic activity of gut microbiota with performance. Under this paradigm, the composition, and periodisation of diet and supplementation, if necessary, during a Grand Tour should be designed, not only from a physiological or metabolic perspective, but also to modulate the composition of the gut microbiota towards that which can allow better performance at each moment of the competition, depending on the accumulated loads.

## Data Availability

All data produced in the present study are available upon reasonable request to the authors.

https://www.ncbi.nlm.nih.gov/bioproject/PRJNA645285

## ACKNOWLEDGEMENTS

Authors wish to thank funding from FICYT-Government of the Principality of Asturias (Ayudas para grupos de investigación del Principado de Asturias, grant AYUD/2021/51347) and Spanish Ministry of Science and Innovation (MICINN, grant PID2021-127812OB-I00), to FL. We also acknowledge the funding of the grant from the Spanish State Research Agency RTI2018-095021-J-I00 (funded by (MCIU/AEI/FEDER, UE), to LR.

The authors acknowledge the technical support provided by Servicios Científico-Técnicos de la Universidad de Oviedo.

## Notes

### Competing Interest Statement

The authors have declared no competing interest.

### Funding Statement

This study was funded by FICYT-Government of the Principality of Asturias (Ayudas para grupos de investigacion del Principado de Asturias, grant AYUD/2021/51347) and Spanish Ministry of Science and Innovation (MICINN, grant PID2021-127812OB-I00), to FL. We also acknowledge the funding of the grant from the Spanish State Research Agency RTI2018-095021-J-I00 (funded by (MCIU/AEI/FEDER, UE), to LR.

### Author Declarations

Both the study design and the informed consent were reviewed and authorized by the Principality of Asturias Research Ethics Committee, Spain (Ref.: 238/19).

## REFERENCES

1. Qin J, Li Y, Cai Z, et al. A metagenome-wide association study of gut microbiota in type 2 diabetes. Nature 2012;490(7418):55–60. doi: 10.1038/nature11450 [published Online First: 2012/10/02]

2. Tang WH, Kitai T, Hazen SL. Gut Microbiota in Cardiovascular Health and Disease. Circ Res 2017;120(7):1183–96. doi: 10.1161/CIRCRESAHA.117.309715 [published Online First: 2017/04/01]

3. Hawley JA, Hargreaves M, Joyner MJ, et al. Integrative biology of exercise. Cell 2014;159(4):738–49. doi: 10.1016/j.cell.2014.10.029

4. Bermon S, Petriz B, Kajeniene A, et al. The microbiota: an exercise immunology perspective. Exerc Immunol Rev 2015;21:70–9. [published Online First: 2015/04/01]

5. Gilbert JA, Blaser MJ, Caporaso JG, et al. Current understanding of the human microbiome. Nat Med 2018;24(4):392–400. doi: 10.1038/nm.4517 [published Online First: 2018/04/11]

6. Allen JM, Berg Miller ME, Pence BD, et al. Voluntary and forced exercise differentially alters the gut microbiome in C57BL/6J mice. J Appl Physiol (1985) 2015;118(8):1059–66. doi: 10.1152/japplphysiol.01077.2014 [published Online First: 2015/02/14]

7. Allen JM, Mailing LJ, Cohrs J, et al. Exercise training-induced modification of the gut microbiota persists after microbiota colonization and attenuates the response to chemically-induced colitis in gnotobiotic mice. Gut Microbes 2018;9(2):115–30. doi: 10.1080/19490976.2017.1372077 [published Online First: 2017/09/02]

8. Barton W, Penney NC, Cronin O, et al. The microbiome of professional athletes differs from that of more sedentary subjects in composition and particularly at the functional metabolic level. Gut 2018;67(4):625–33. doi: 10.1136/gutjnl-2016-313627 [published Online First: 2017/04/01]

9. Fernandez J, Fernandez-Sanjurjo M, Iglesias-Gutierrez E, et al. Resistance and Endurance Exercise Training Induce Differential Changes in Gut Microbiota Composition in Murine Models. Front Physiol 2021;12:748854. doi: 10.3389/fphys.2021.748854 [published Online First: 20211224]

10. Okamoto T, Morino K, Ugi S, et al. Microbiome potentiates endurance exercise through intestinal acetate production. Am J Physiol Endocrinol Metab 2019;316(5):E956–E66. doi: 10.1152/ajpendo.00510.2018 [published Online First: 2019/03/13]

11. Scheiman J, Luber JM, Chavkin TA, et al. Meta-omics analysis of elite athletes identifies a performance-enhancing microbe that functions via lactate metabolism. Nat Med 2019;25(7):1104–09. doi: 10.1038/s41591-019-0485-4 [published Online First: 2019/06/27]

12. Hsu YJ, Chiu CC, Li YP, et al. Effect of intestinal microbiota on exercise performance in mice. J Strength Cond Res 2015;29(2):552–8. doi: 10.1519/JSC.0000000000000644 [published Online First: 2014/08/22]

13. Maughan RJ, Shirreffs SM, Vernec A. Making Decisions About Supplement Use. Int J Sport Nutr Exerc Metab 2018;28(2):212–19. doi: 10.1123/ijsnem.2018-0009 [published Online First: 2018/03/23]

14. Donati Zeppa S, Agostini D, Gervasi M, et al. Mutual Interactions among Exercise, Sport Supplements and Microbiota. Nutrients 2019;12(1) doi: 10.3390/nu12010017 [published Online First: 2019/12/22]

15. Chicharro JL, Hoyos J, Bandres F, et al. Thyroid hormone levels during a 3-week professional road cycling competition. Horm Res 2001;56(5-6):159–64. doi: 10.1159/000048112 [published Online First: 2002/03/23]

16. Lucia A, Diaz B, Hoyos J, et al. Hormone levels of world class cyclists during the Tour of Spain stage race. Br J Sports Med 2001;35(6):424–30. doi: 10.1136/bjsm.35.6.424 [published Online First: 2001/12/01]

17. Yang W, Liu Y, Yang G, et al. Moderate-Intensity Physical Exercise Affects the Exercise Performance and Gut Microbiota of Mice. Front Cell Infect Microbiol 2021;11:712381. doi: 10.3389/fcimb.2021.712381 [published Online First: 20210924]

18. Moris G, Arboleya S, Mancabelli L, et al. Fecal microbiota profile in a group of myasthenia gravis patients. Sci Rep 2018;8(1):14384. doi: 10.1038/s41598-018-32700-y [published Online First: 2018/09/28]

19. Borg G. Psychophysical scaling with applications in physical work and the perception of exertion. Scand J Work Environ Health 1990;16 Suppl 1:55–8. doi: 10.5271/sjweh.1815 [published Online First: 1990/01/01]

20. Kentta G, Hassmen P. Overtraining and recovery. A conceptual model. Sports Med 1998;26(1):1–16. doi: 10.2165/00007256-199826010-00001 [published Online First: 1998/09/18]

21. Rodriguez IT, Ballart JF, Pastor GC, et al. [Validation of a short questionnaire on frequency of dietary intake: reproducibility and validity]. Nutr Hosp 2008;23(3):242–52. [published Online First: 2008/06/19]

22. Garcia-Roves PM, Terrados N, Fernandez S, et al. Comparison of dietary intake and eating behavior of professional road cyclists during training and competition. Int J Sport Nutr Exerc Metab 2000;10(1):82–98. doi: 10.1123/ijsnem.10.1.82 [published Online First: 2000/08/12]

23. Rodriguez-Alonso M, Fernández-García B. Evolution of the use of sports supplements. PharmaNutrition 2020;14:100239. doi: https://doi.org/10.1016/j.phanu.2020.100239

24. Hughes RL. A Review of the Role of the Gut Microbiome in Personalized Sports Nutrition. Front Nutr 2019;6:191. doi: 10.3389/fnut.2019.00191 [published Online First: 20200110]

25. Bongiovanni T, Yin MOL, Heaney LM. The Athlete and Gut Microbiome: Short-chain Fatty Acids as Potential Ergogenic Aids for Exercise and Training. Int J Sports Med 2021;42(13):1143–58. doi: 10.1055/a-1524-2095 [published Online First: 20210713]

26. Morrison DJ, Preston T. Formation of short chain fatty acids by the gut microbiota and their impact on human metabolism. Gut Microbes 2016;7(3):189–200. doi: 10.1080/19490976.2015.1134082 [published Online First: 20160310]

27. Luo L, Li R, Wang G, et al. Age-dependent effects of a high-fat diet combined with dietary advanced glycation end products on cognitive function and protection with voluntary exercise. Food Funct 2022;13(8):4445–58. doi: 10.1039/d1fo03241k [published Online First: 20220420]

28. Lamoureux EV, Grandy SA, Langille MGI. Moderate Exercise Has Limited but Distinguishable Effects on the Mouse Microbiome. mSystems 2017;2(4) doi: 10.1128/mSystems.00006-17 [published Online First: 20170822]

29. Zhao X, Zhang Z, Hu B, et al. Response of Gut Microbiota to Metabolite Changes Induced by Endurance Exercise. Front Microbiol 2018;9:765. doi: 10.3389/fmicb.2018.00765 [published Online First: 20180420]

30. Jager R, Purpura M, Stone JD, et al. Probiotic Streptococcus thermophilus FP4 and Bifidobacterium breve BR03 Supplementation Attenuates Performance and Range-of-Motion Decrements Following Muscle Damaging Exercise. Nutrients 2016;8(10) doi: 10.3390/nu8100642 [published Online First: 20161014]

31. Huang WC, Hsu YJ, Huang CC, et al. Exercise Training Combined with Bifidobacterium longum OLP-01 Supplementation Improves Exercise Physiological Adaption and Performance. Nutrients 2020;12(4) doi: 10.3390/nu12041145 [published Online First: 20200419]

32. Estaki M, Pither J, Baumeister P, et al. Cardiorespiratory fitness as a predictor of intestinal microbial diversity and distinct metagenomic functions. Microbiome 2016;4(1):42. doi: 10.1186/s40168-016-0189-7 [published Online First: 20160808]

33. Resende AS, Leite GSF, Lancha Junior AH. Changes in the Gut Bacteria Composition of Healthy Men with the Same Nutritional Profile Undergoing 10-Week Aerobic Exercise Training: A Randomized Controlled Trial. Nutrients 2021;13(8) doi: 10.3390/nu13082839 [published Online First: 20210818]

34. Portincasa P, Bonfrate L, Vacca M, et al. Gut Microbiota and Short Chain Fatty Acids: Implications in Glucose Homeostasis. Int J Mol Sci 2022;23(3) doi: 10.3390/ijms23031105 [published Online First: 20220120]

35. Deleu S, Machiels K, Raes J, et al. Short chain fatty acids and its producing organisms: An overlooked therapy for IBD? EBioMedicine 2021;66:103293. doi: 10.1016/j.ebiom.2021.103293 [published Online First: 20210401]

36. Frampton J, Murphy KG, Frost G, et al. Short-chain fatty acids as potential regulators of skeletal muscle metabolism and function. Nat Metab 2020;2(9):840–48. doi: 10.1038/s42255-020-0188-7 [published Online First: 20200330]

37. Rios-Covian D, Ruas-Madiedo P, Margolles A, et al. Intestinal Short Chain Fatty Acids and their Link with Diet and Human Health. Front Microbiol 2016;7:185. doi: 10.3389/fmicb.2016.00185 [published Online First: 20160217]

38. Dziewiecka H, Buttar HS, Kasperska A, et al. Physical activity induced alterations of gut microbiota in humans: a systematic review. BMC Sports Sci Med Rehabil 2022;14(1):122. doi: 10.1186/s13102-022-00513-2 [published Online First: 20220707]

39. D’Souza G, Shitut S, Preussger D, et al. Ecology and evolution of metabolic cross-feeding interactions in bacteria. Nat Prod Rep 2018;35(5):455–88. doi: 10.1039/c8np00009c [published Online First: 20180525]

40. Vacca M, Celano G, Calabrese FM, et al. The Controversial Role of Human Gut Lachnospiraceae. Microorganisms 2020;8(4) doi: 10.3390/microorganisms8040573 [published Online First: 20200415]

41. Jeukendrup AE. Training the Gut for Athletes. Sports Med 2017;47(Suppl 1):101–10. doi: 10.1007/s40279-017-0690-6

42. Huang WC, Chen YH, Chuang HL, et al. Investigation of the Effects of Microbiota on Exercise Physiological Adaption, Performance, and Energy Utilization Using a Gnotobiotic Animal Model. Front Microbiol 2019;10:1906. doi: 10.3389/fmicb.2019.01906 [published Online First: 20190820]

