## Supplemental Figures for "Dynamics of gut microbiota and short-chain fatty acids during a cycling Grand Tour are related to exercise performance and modulated by dietary intake"

### Slide 1
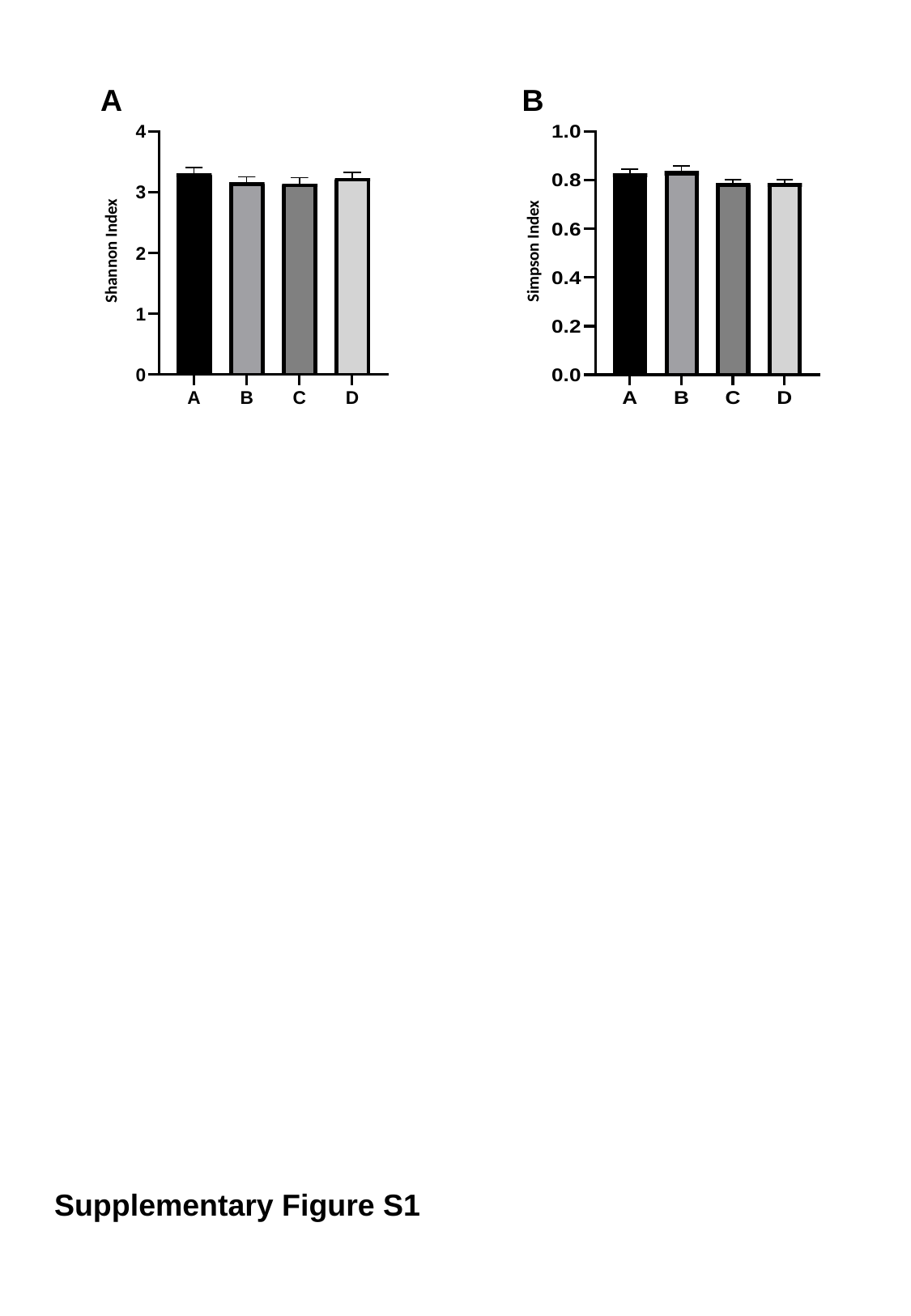

A
B
Shannon Index
Simpson Index
Supplementary Figure S1

### Slide 2
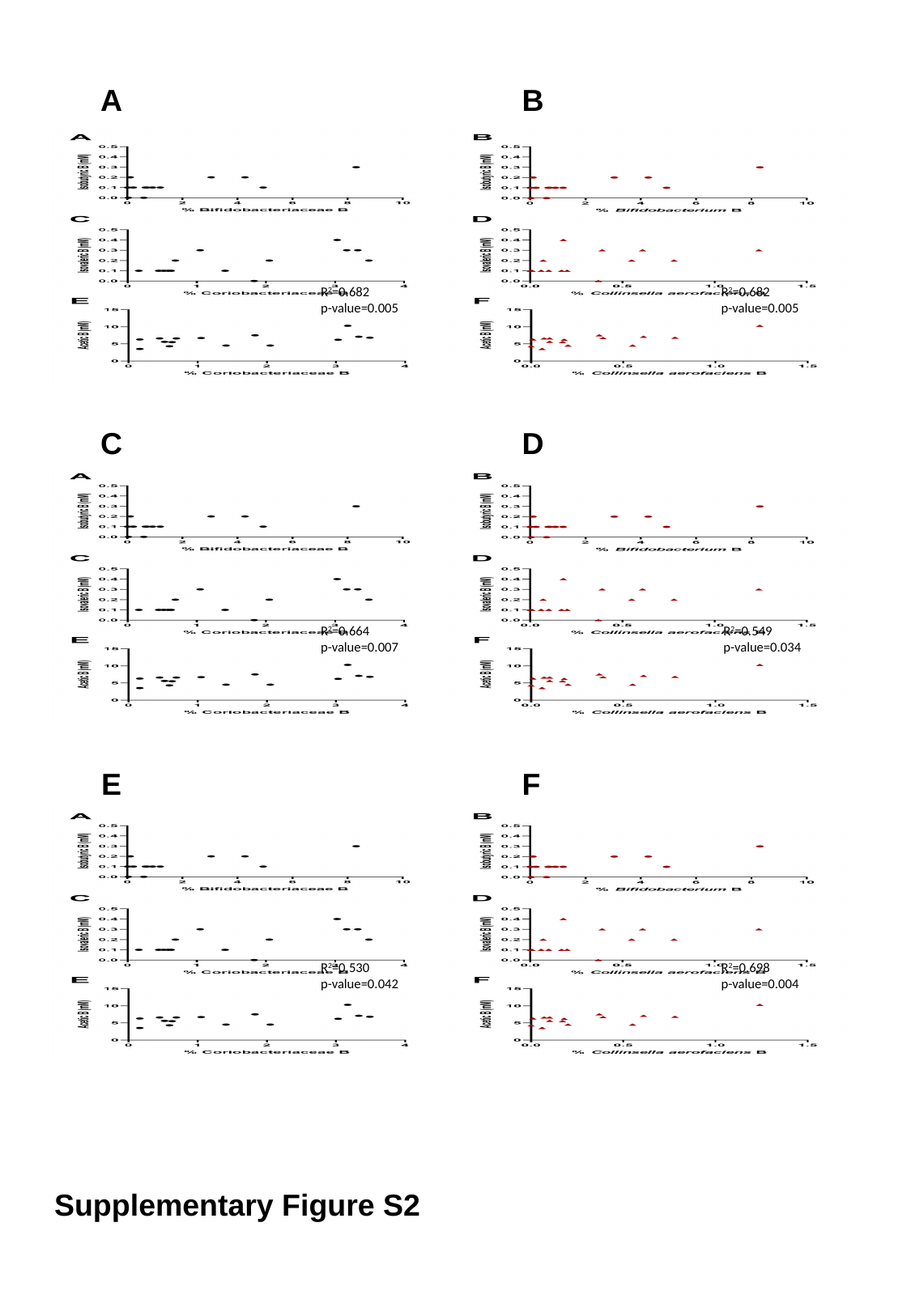

A
B
R^2^=0.682
p-value=0.005
R^2^=0.682
p-value=0.005
C
D
R^2^=0.549
p-value=0.034
R^2^=0.664
p-value=0.007
E
F
R^2^=0.698
p-value=0.004
R^2^=0.530
p-value=0.042
Supplementary Figure S2

### Slide 3
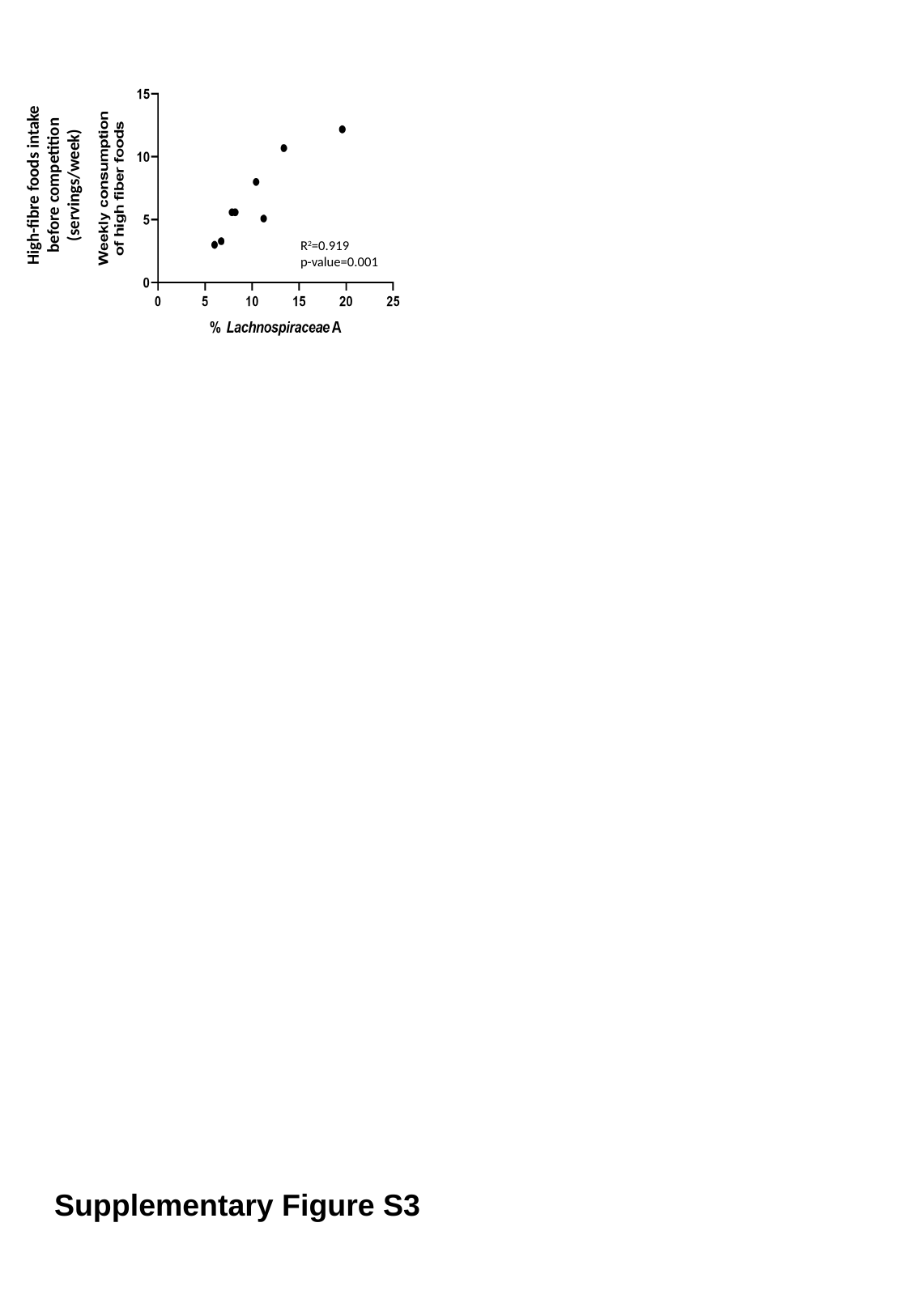

High-fibre foods intake before competition (servings/week)
R^2^=0.919
p-value=0.001
Supplementary Figure S3
